# Validity and reliability of the Hypersensitive Narcissism Scale (HSNS) in China

**DOI:** 10.1101/2024.09.28.24312621

**Authors:** Yuxin Li, Fan Zou, Hao Cheng

## Abstract

**Background:** Wink distinguishes between explicit and implicit narcissism, the latter being vulnerability-sensitive narcissism, which manifests itself as outward humility and inhibition masking an underlying pompous outlook and sense of entitlement. Early measures of vulnerability-sensitive narcissism used Murray’s Narcissism Scale, which was explored by measuring the narcissism levels of Harvard undergraduates. Hendin and Cheek subsequently developed the Vulnerable Sensitive Narcissism Scale based on this narcissism scale. In addition to this, there are few ways to measure vulnerability-sensitive narcissism, and Chinese measurement tools on narcissism mainly measure explicit narcissism, neglecting to accurately measure vulnerability-sensitive narcissism, and failing to completely respond to an individual’s implicit narcissistic traits.

**Aim:** The hypersensitive narcissism scale was applied to college students to explore the factor structure of the scale under the background of Chinese culture, and to test its reliability and validity.

**Methods:** 1000 questionnaires were distributed to college students, and 912 valid questionnaires were collected. Self-Esteem Scale, Aggression Scale and Narcissism subscale of Dark Triad Scale were used as the criterion. After 3 weeks, 100 college students were randomly selected for retest.

**Results:** The exploratory factor analysis showed that one dimension should be extracted. Eigenvalue was 5.352 and 53.518% cumulative variance interpretation rate. The confirmatory factor analysis showed that the scale structure validity was good (χ^*2*^*/df*=3.237,CFI=0.936,TLI=0.931, RMSEA=0.066,SRMR=0.053). Internal consistency reliability of the scale was 0.785; The test-retest reliability was 0.833, and the criterion correlation validity reached a significant level.

**Conclusion:** The Chinese version of hypersensitive narcissism scale has good reliability and validity, which can effectively measure hypersensitive narcissism. The localisation of the scale can better serve the research and development of the university student population on the one hand, and on the other hand, it can help the research in this field to have a more in-depth development in China.

## 1 Introduction

Narcissism is defined as a pervasive sense of grandiosity, self-focus, and privilege accompanied by a strong desire to be recognized and appreciated by others ^[1]^. Wink ^[2]^ distinguishes narcissism into explicit and implicit narcissism based on the psychodynamic theory proposed by Kernberg ^[3]^. He referred to implicit narcissism as vulnerable, sensitive narcissism and explicit narcissism as pompous, performance narcissism. He argued that the two types of narcissism are measurably differentiated, and not correlated.

Hypersensitive narcissism masks an underlying pompous outlook and sense of entitlement through outward humility and inhibition. Today more and more researchers are devoting their energies to the psychological and behavioral effects of Hypersensitive narcissism on individuals ^[4]^. It has been shown that vulnerability-sensitive narcissism is a risk factor for self-objectification and selfbody imagery^[5]^ ; vulnerability-sensitive narcissism is a major predictive variable for depression and is positively correlated with the degree of depression in depressed individuals^[6-7]^ ; hypersensitive narcissism is also an effective predictor of narcissism levels in patients in treatment for substance abuse or dependence^[8]^ . Meanwhile, Stone et al. pointed out that rejection sensitivity is the most fundamental difference between sensitive narcissism and flamboyant expressive narcissism^[9]^, which is manifested in a strong sensitivity to and fear of social rejection, which causes people to constantly experience high levels of rumination and avoid criticism and evaluation. At the same time, hypersensitive narcissists exhibit social inhibition and are often isolated or lonely^[10]^. Excessive levels of vulnerability sensitivity in social relationships raise the risk of cardiovascular disease^[11]^.

Early measures of vulnerability-sensitive narcissism utilized Murray ‘s Narcissism Scale^[4]^, which was derived by measuring the narcissism levels of Harvard undergraduates. Subsequently Hendin and Cheek^[12]^ developed the Hypersensitive Narcissism Scale (HSNS) based on Murray ‘s Narcissism Scale^[4]^, which consists of 10 questions with a single dimension. It measures a more defensive and insecure type of narcissism that is associated with low self-esteem and shame^[13]^. In addition to this, there are few ways to measure vulnerability-sensitive narcissism, and domestic instruments on narcissism mainly measure explicit narcissism, neglecting to accurately measure vulnerability-sensitive narcissism and failing to completely respond to the individual’s implicit narcissistic traits. To summarize, more and more studies focus not only on the pompous and expressive side of narcissism, but also on its fragile and sensitive side. The compilation of the HSNS effectively solves the problem of measuring hypersensitive narcissism, but it lacks the exploration of its factor structure in the context of the Chinese culture; at the same time, college students, as an important period of time before entering the society, the fragile and sensitive narcissism will have a great impact on the growth and development of Chinese college students, and should be cultivated in all aspects to improve their own development. In this context, the localization of the scale in China is particularly important. In this context, it is important to localize the scale in China. The present study was conducted to revise the scale for college students and to examine the reliability of the HSNS in the Chinese cultural context. It is hoped that this study will provide a valid and reliable assessment tool for the study of implicit narcissism in China and enrich the research results in the field of narcissism.

## 2 Materials and Method

### 2.1 Participants

We randomly recruited 1000 college students in Northeast Normal University(Jilin Province, China)and Harbin Normal University(Heilongjiang Province, China) by online survey. Sample 1(N=600) was used for exploratory factor analysis(EFA), which consisted of 559 valid questionnaires. Sample 2(N=400) was used for confirmatory factor analysis(CFA), which consisted of 353 valid questionnaires. Exploratory questionnaires are more than confirmatory questionnaires to ensure the stability and reliability of the results. Excluding 88 invalid questionnaires (regular answers), 912 valid questionnaires remained, with an effective rate of 91.2%. The final sample consisted of 220 males and 692 females, aged from 18 to 24 (M=20.72,SD=2.22). 37.6% were from urban and 62.4% were from rural;42.1% were Only Children and 57.9% were None-Only Children.

After three weeks of the data recovery, 100 college students were randomly selected for retest. The retest sample consisted of 97 data (30 males and 67 females) after excluding invalid questionnaires.

### 2.2 Instruments

#### 2.2.1 Hypersensitivity Narcissism Scale(HSNS)

The Hypersensitivity Narcissism Scale was developed by Hendin and Cheek^[5]^ with reference to Murray’s Narcissistic Scale^[4]^. Assess the extent of an individual’s covert narcissism. It has a single dimension with 10 items. Participants answering on a five-point Likert-type scale, ranging from 1 =“very nontypical” to 5 = “very typical “.The higher total score, the higher the degree of covert narcissism. With the authorization of the author of HSNS, the original scale is translated and translated back, according to Chinese expression habits, under the premise of ensuring that the meaning of the topic remains unchanged, it is easier to be understood and expressed more accurately by Chinese college students. Finally, after the advice and modification of the language expression and professional knowledge field of psychologists, the Chinese version of the hypersensitivity narcissistic scale was formed.

#### 2.2.2 Self-Esteem Scale

The Self-Esteem Scale uses the classic Rosenberg’s Self-Esteem Scale^[14]^, it is a single dimension including 10 items, of which 5 are inversely scored. Participants answering on a four-point Likert-type scale, ranging from 1 =“very inconsistent” to 4 = “very compliant”. The reverse scoring questions are converted and added, and the higher the total score, the higher the individual’s self-esteem. The Chinese version of the Self-esteem Scale has good reliability and validity^[15]^. In this study, the Cronbach’s α of this scale was 0.836.

#### 2.2.3 Buss-Perry Aggression Scale (BPAS)

BPAS was developed by Buss and Perry^[16]^. The Chinese version of BPAS was revised by Lv et al.^[17]^with four subscales(hostility, impulsivity, physical aggression, and irritability),totaling 22 items. Participants answering on a five-point Likert-type scale(e.g., 1=“Strongly Disagree” to 5=“Strongly Agree”).The higher the mean score, the higher levels of aggression. In this survey, the Cronbach’s α of this scale was 0.894, and the Cronbach’s α of the four subscales were from 0.674 to 0.852.

#### 2.2.4 Narcissism of the Dark Triad Scale

The Dark Triad Scale was compiled by Paulhus and Williams^[18]^, including three dimensions of Machiavellianism, narcissism, and psychopathy. The Chinese version of the narcissistic subscale revised by Geng et al^[19]^. Participants answering 9 items on a five-point Likert-type scale(e.g., 1=“Strongly Disagree” to 5=“Strongly Agree”), of which 3 are scored in reverse. The reverse scoring questions are converted and added, and the higher the total score, the higher the degree of individual narcissism. The Cronbach’s α of this scale is 0.754.

### 2.3 Data Analysis

This study adopted descriptive statistics, item analysis, correlation analysis, variance analysis and exploratory factor analysis(EFA) via SPSS 25.0, and confirmatory factor analysis (CFA) via Mplus 7.4.

## 3 Results

### 3.1 Item Analysis

Item analysis was used to calculate the discrimination of each item. Firstly, each item’s item-total correlation was between 0.369 and 0.583, which met the criterion(greater than 0.30). Additionally, we selected the highest 27% as the high-score group and the lowest 27% as the low-score group to conduct a t-test to ensure that the two groups could be differentiated. The results showed that the difference between the two groups was significant(p<0.01).

### 3.2 Construct Validity

#### 3.2.1 Exploratory Factor Analysis

After item analysis met the criterion, principal component analysis and Promax oblique rotation were adopted to EFA of the questionnaires(N=559).Firstly, the Kaiser-Meyer-Olkin(KMO) value was 0.887, and the results of Bartlett’s sphericity test showed χ^*2*^*/df* =1868.735,*df*=45,*P*<0.001, indicating that it was suitable for EFA. Secondly, the single factor with an eigenvalue higher than 1 was obtained by principal component analysis and oblique rotation. The eigenvalue was 5.352, and the total interpretation rate of variance was 53.518%, suggesting a single dominant factor is suitable. The scree plot also implied that the one-factor structure was acceptable. It is consistent with the original scale. Loadings of the items are all greater than 0.6, from 0.643 to 0.814(Table 1).

**Table 1.** Results of Factor Loadings of HSNS(sample1 N=559)

| HSNS |  |  |  |  |  |
| --- | --- | --- | --- | --- | --- |
| Item | F | Item | F | Item | F |
| 9 | 0.814 | 8 | 0.765 | 1 | 0.689 |
| 6 | 0.803 | 3 | 0.751 | 7 | 0.643 |
| 4 | 0.795 | 10 | 0.735 |  |  |
| 2 | 0.788 | 5 | 0.703 |  |  |

Note: F = Factor Loading

#### 3.2.2 Confirmatory Factor Analysis

The result of CFA of Sample2(N=353) in Mplus7.4 showed that the fitting results all met the statistical standards: χ^*2*^*/df*=3.237, CFI=0.936, TLI=0.931, RMSEA=0.066, SRMR=0.053, indicating that the one-factor structure of the Chinese version of the HSNS was fitting well.

#### 3.2.3 Criterion-related Validity

Pearson correlation analysis was used to analyze the criterion-related validity(Table 2).The results showed that hypersensitivity narcissism was significantly negatively correlated with self-esteem and extremely positively correlated with aggression and narcissism.

**Table 2.** Correlation between HSNS and Criterions(sample1,sample2 N=912)

|  | HSNS | Self-esteem | Aggression | Narcissism |
| --- | --- | --- | --- | --- |
| HSNS | 1 |  |  |  |
| Self-esteem | -0.187** | 1 |  |  |
| Aggression | 0.543** | -0.388** | 1 |  |
| Narcissism | 0.157** | 0.188** | 0.230** | 1 |

Note : \* $P < 0.05$ , \*\* $P < 0.01$

### 3.3 Reliability Analysis

Cronbach’s α coefficient for Sample1 and Sample2 was 0.785, with 912 valid data. The retest reliability coefficient was 0.833. These coefficient results indicate that the reliability of Chinese HSNS is good.

### 3.4 Analysis of demographic variables

First, we conduct t-test for the gender, the place of origin and whether the Only-children of HSNS separately, and the results were shown in Table 3 to 5.

**Table 3.** Variance analysis of gender in HSNS(N=912, M±SD)

| | male | female | *t* | 1-β | *d* |
| --- | --- | --- | --- | --- | --- |
| HSNS | 2.966±0.5<br>55 | 2.987±0.5<br>14 | -0.509 | 0.080 | 0.039 |

**Table 4.** Variance analysis of student’s origin in HSNS(N=912, M±SD)

| | urban | rural | *t* | 1-β | *d* |
| --- | --- | --- | --- | --- | --- |
| HSNS | 3.089±0.5<br>54 | 2.917±0.4<br>95 | 4.874** | 0.998 | 0.331 |

Note : \* $P$ <0.05 , \*\* $P$ <0.01

**Table 5.** Variance analysis of Whether Only-Children differences in HSNS (N=912, M±SD)

| | Only-Children | Non-Only | *t* | 1-β | *d* |
| --- | --- | --- | --- | --- | --- |
| HSNS | 3.062±0.5<br>38 | 2.923±0.5<br>06 | 3.995** | 0.979 | 0.268 |

Note : \* $P$ <0.05 , \*\* $P$ <0.01

These results showed that the difference between sexes in Hypersensitive narcissism was not significant, and the overall average score of girls was slightly higher than that of boys. The difference in the place of origin of students is extremely significant, and the overall average score of urban students is significantly higher than that of rural students; The difference in whether or not the Only-children is extremely significant, and the average score of Only-Children students is significantly higher than that of Non-Only children.

Secondly, the analysis of variance is performed on the academic performance, and the results are shown in Table 6.

**Table 6.**
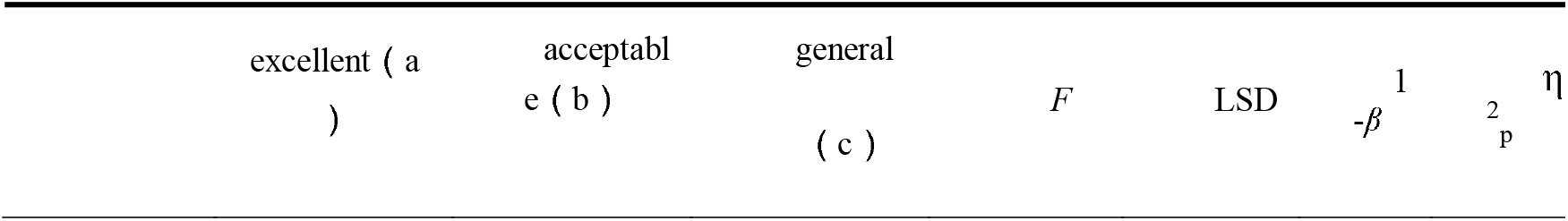
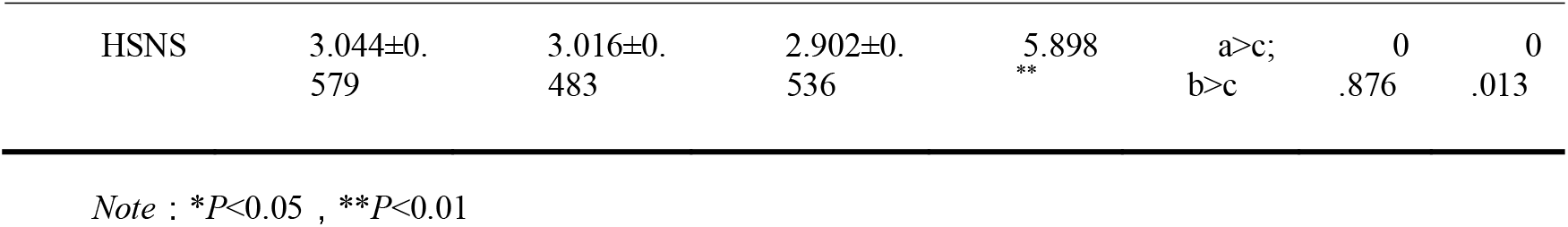
Variance analysis of academic performance in HSNS(N=912, M±SD)

According to the post-mortem LSD method, the Hypersensitive narcissism of students with excellent academic performance is significantly higher than that of students with general academic performance, and similarly, the Hypersensitive narcissism of students with acceptable academic performance is also extremely significantly higher than that of students with general academic performance.

## 4 Discussion

Wink[2] pioneered research in the field of narcissism by distinguishing between explicit and implicit narcissism, contributing to the development of implicit narcissism, and the Vulnerable Sensitive Narcissism Scale measures narcissism as one of the many facets of a higher structure proposed by Maslow[20] to view this narcissism as psychological insecurity. And the development of the Vulnerable Sensitive Narcissism Scale by Hendin and Cheek^[12]^ has allowed for the valid measurement of implicit narcissism. The Vulnerability Sensitive Narcissism Scale was revised in order to be able to find a rational structure and suitable expression in the Chinese context.

The accuracy of the topic translation was ensured by English-Chinese translation and back translation. With the upper and lower 27% as the grouping criterion, a difference test was conducted for the 10 question items in the high and low groupings, and all the questions reached the significant level, which indicated that each question item was reasonably set up with a good level of differentiation. 600 questionnaires were distributed as exploratory factor analysis data, and 400 questionnaires were used as validation factor analysis, unlike previous factor analysis conducted in half, more data for exploration can get more stable and accurate scale structure^[21-22]^ . Psychological concepts do not exist in a perfectly orthogonal situation, so a more realistic oblique rotation was used for the analysis, and with the KMO value and Bartlett’s spherical test satisfied, a one-factor, 10-item Chinese version of the Vulnerable Sensitive Narcissism Scale was obtained, with the same structure as the original scale. The covariance, cumulative variance explained also satisfied the measurement criteria. Based on the explored scale structure was validated with another batch of data, and the structural validity of the scale was judged based on the degree of data fit. All the fit indicators met the criteria, and the factor loadings of the 10 question items were all above 0.643, that is, the explanatory rates were all above 41%, and each question was representative. The total question correlations of the scale were all greater than 0.3, and the Cronbach’s alpha coefficients were also greater than the psychometric criterion of 0.7. After an interval of 3 weeks, 100 students who had previously taken the questionnaire were randomly selected for retesting, and the retest reliability was 0.833, indicating that the scale was stable across time. Validity scale correlation validity analysis found that fragile sensitive narcissism was significantly negatively correlated with self-esteem, and implicit narcissism is the fragile, sensitive part of narcissism, and there is research support that narcissism is associated with low self-esteem^[13]^. Vulnerable sensitive narcissism was significantly and positively correlated with aggression, with some research suggesting that when a narcissist’s view of self is challenged and questioned, the instability of self-esteem and uncertainty of self-worth bring about an overall sensitivity that makes the narcissist inclined to use aggression in response^[23]^. Vulnerable and sensitive narcissism was also significantly and positively correlated with the narcissism subscale of the Dark Triad Scale, which measures a predominantly antisocial personality trait, which coincides with the vulnerability and sensitivity of covert narcissism.

The analysis of demographic variables revealed that the difference between men and women in vulnerability-sensitive narcissism was not significant, but since the sample was selected from two teacher training colleges and the ratio of men to women was more than 1:3, it could not be inferred that female students had higher levels of covert narcissism than their male counterparts. The level of narcissism of urban students is significantly higher than that of rural students. Urban students face more competitive pressure than rural students since childhood, which makes them sensitive and uneasy. The level of narcissism of only-child students is significantly higher than that of non-only-child students, mainly due to the psychological tendency of only-child students to be more criticized and ignored by their parents, which gradually leads to their fragile and sensitive narcissism^[24]^. Students with good grades and those with fair grades were significantly higher than those with average grades, with those with good grades scoring higher in implicit narcissism. Vulnerable and sensitive narcissists are more sensitive to aggression and provocation and are more likely to compete than others^[25]^. So while avoiding aggressive competition, you can prove that you are better than others through good academic performance, so as to be recognized and at the same time avoid feeling uneasy.

## 5 Limitation

This study still has some limitations. First, in terms of subject selection, the study only conducted a questionnaire survey for college students, which may have limited representativeness and could not comprehensively reflect the vulnerability-sensitive narcissistic characteristics of people of different ages, occupations, and cultural backgrounds. Moreover, the samples mainly came from Northeast Normal University and Harbin Normal University, which are geographically more concentrated, and there may be some regional cultural bias, and subsequent studies can further verify the applicability to more age groups or special groups (e.g., clinical patients, corporate employees, etc.); second, in terms of the questionnaire administration, it will inevitably produce the social approval effect as well as self-report bias, and experimental method or Finally, as international scholars deepen their research on the measurement dimensions and accuracy of the HSNS, the specific relationship and mechanism of the HSNS with other psychological factors (e.g., anxiety, depression, etc.) in the Chinese cultural context can be explored in depth.

## 6 Implication

This study is the first to explore the characteristics of Hypersensitive narcissism in Chinese college students and to revise the English version of the HSNS into Chinese. Extending the applicability of the HSNS to Chinese culture could facilitate cross-cultural comparisons of covert narcissism in future studies. The results indicate that the Chinese version of the HSNS scale has good validity and reliability and provides a valid measurement tool for screening covert narcissism in the Chinese context.

## Data Availability

All data produced in the present study are available upon reasonable request to the authors.
All data are contained in the manuscript.

